# Epidemiological Analysis of HIV and HPgV Co-infection in a Brazilian Population

**DOI:** 10.1101/2024.08.27.24312685

**Authors:** Evônio de Barros Campelo Júnior, Ana Maria Passos de Castilho, Viviane Martha Santos de Morais, Georgea Gertrudes de Oliveira Mendes Cahú, Gabriel Galindo Cunha, Vitoria Maria de Araújo Oliveira, Ricardo de Arraes Alencar Ximenes, Maria Rosângela Cunha Duarte Coêlho

**Author notes:** Corresponding author: Maria Rosângela Cunha Duarte Coêlho. **Financing:** This work did not receive any specific grant from any funding agency. **Consent to publish:** We declare that consent for publication has been obtained.

## Abstract

**Objectives:** This study aimed to determine the prevalence of Human Pegivirus 1 infection among people living with HIV (PLWHIV) in Northeast Brazil and identify associated risk factors, given the potential impact of HPgV on HIV disease progression

**Methods:** We conducted a cross-sectional analysis of 249 PLWHIV receiving care at a tertiary hospital in Northeast Brazil. Bivariate and multivariate logistic regression models were used to assess the association between sociodemographic characteristics, risk behaviors, healthcare access, and HPgV infection.

**Results:** The prevalence of HPgV infection among the study population was 23.3%. Inhaled drug use emerged as a significant risk factor for HPgV infection in the multivariate analysis.

**Conclusions:** A concerning prevalence of HPgV infection exists among PLWHIV in Northeast Brazil. The identification of inhaled drug use as a risk factor suggests potential transmission routes beyond traditional modes. These findings highlight the need for targeted public health interventions and harm reduction strategies to address this significant public health concern.

## INTRODUCTION

The Human Pegivirus 1 (HPgV-1) is a single-stranded, positive-sense RNA virus belonging to the Flaviviridae family, with a genome comprising approximately 9400 nucleotides (1; 2). Although HPgV is not associated with specific diseases, research suggests a correlation between CD4+ lymphocyte count and human immunodeficiency virus (HIV) infection, which may influence the progression of HIV infection in co-infected individuals. In fact, some studies have found that HPgV’s viral influence on lymphocyte counts is linked to a slower progression of HIV infection in co-infected individuals (3; 4), and co-infected patients have been shown to experience a decrease in morbidity and mortality (3; 5). Furthermore, prior HPgV infection of lymphocytes can inhibit HIV replication, which may contribute to the observed slower progression of HIV infection. (1).

Researchers have analyzed the prevalence of HPgV among people living with HIV/AIDS (PLWHA) in several studies (6; 7; 8), and found that it varies across regions of the world. Notably, they have observed an inverse relationship between the prevalence and nd the level of regional development (9; 10; 11). Studies have shown that the overall prevalence of HPgV-1 monoinfection ranges from 1% to 4% in North America and Europe, whereas it ranges from 5% to 19% in Africa, Asia, and South America (12). In the context of coinfection, researchers have reported high HIV/HPgV prevalence rates in some regions of Brazil, ranging from 17% in the North (13) to 34% in the South (14).

Given the high global prevalence of HPgV infection, we conducted this study to estimate the prevalence of HIV/HPgV co-infection and identify the associated risk factors. Specifically, we sought to investigate the relationship between HPgV infection and HIV disease progression, as well as to determine the demographic and clinical characteristics of individuals co-infected with both viruses. By examining these factors, we aimed to provide a comprehensive understanding of the epidemiology of HIV/HPgV co-infection in a brazilian northeast state, which could inform future research and contribute to the development of more effective prevention and treatment strategies.

## METHODS

### Design and Study Population

In this cross-sectional study, we recruited 249 HIV-positive individuals aged 18 and above who were receiving care at the Department of Infectious and Parasitic Diseases at the Hospital das Clínicas of the Federal University of Pernambuco. During their routine outpatient visits, we invited these patients to participate in our study. Those who provided informed consent were asked to complete a standardized questionnaire developed specifically for this research, which gathered comprehensive data on sociodemographic characteristics, sexual behavior, and potential transmission routes

As part of their routine laboratory examinations at HC-UFPE, we collected blood samples from the participants. These samples were promptly transported to the Laboratory of Virology at the Institute Keizo Asami at the Federal University of Pernambuco, where we processed them to separate plasma and serum, which were then stored at −80°C for future analysis. This project was approved by the Ethics Committee with ID number: 0276.0.172.00-11.

### HPgV RNA Detection

Our HPgV research was conducted in the Clinical Virology Laboratory at the University of São Paulo. We began by extracting RNA from the plasma samples using the Axygen Miniprep Kit, strictly adhering to the manufacturer’s guidelines. Next, we performed the reverse-transcription real-time polymerase chain reaction process using the AgPath-IDTM One-Step RT-PCR Kit, following the method outlined by Alves-Sousa et al. (2012) 15. The reaction mixture included a 5 μL sample, 0.4 μM of both the forward and reverse primers, and 0.15 μM of the probe. The PCR cycling was carried out on an ABI 7500 Real Time PCR System, with an initial cycle at 50 °C for 10 minutes and one cycle at 95 °C for 10 minutes, followed by 45 cycles of 15 seconds at 95 °C and 40 seconds at 60 °C.

### Statistical Analysis

We evaluated the strength and statistical significance of the association between HPgV and HIV infection by calculating odds ratios and confidence intervals, as well as performing chi-square tests or Fisher’s exact tests. To identify variables associated with HPgV infection, we first conducted a bivariate analysis to pinpoint potential variables related to HPgV positivity. In the bivariate analysis, we examined the relationship between various demographic, behavioral, and clinical factors and the presence of HPgV infection. We then selected variables with a p-value less than 0.2 from the bivariate analysis to include in the subsequent multivariate analysis. For the multivariate analysis, we utilized the forward stepwise logistic regression method to develop a predictive model for HPgV presence, allowing us to assess the independent effects of each variable while controlling for potential confounding factors. All the statistic were made by using jamovi software version 2.3.28.

## RESULTS

The analysis revealed a 23.3% prevalence of HPgV RNA among people living with HIV. Detailed insights are provided in Tables 1 and 2, which present the univariate analysis of the association between sociodemographic factors, sexual behavior variables, blood transmission, and HPgV infection in this population.

**Table 1.**
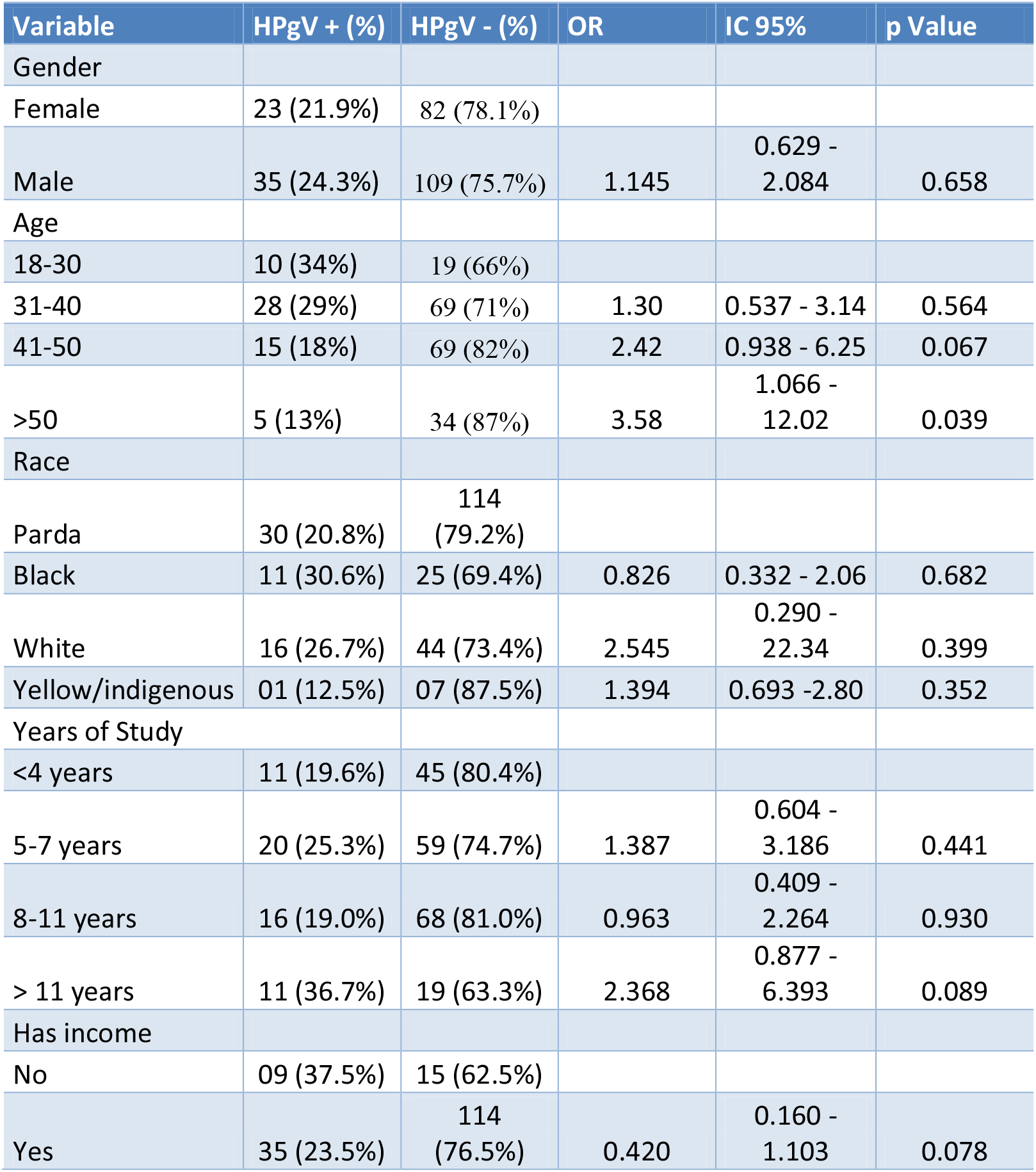
Univariate Analysis of the Association Between Sociodemographic Variables and HPgV Infection in people living with HIV.

**Table 2.**
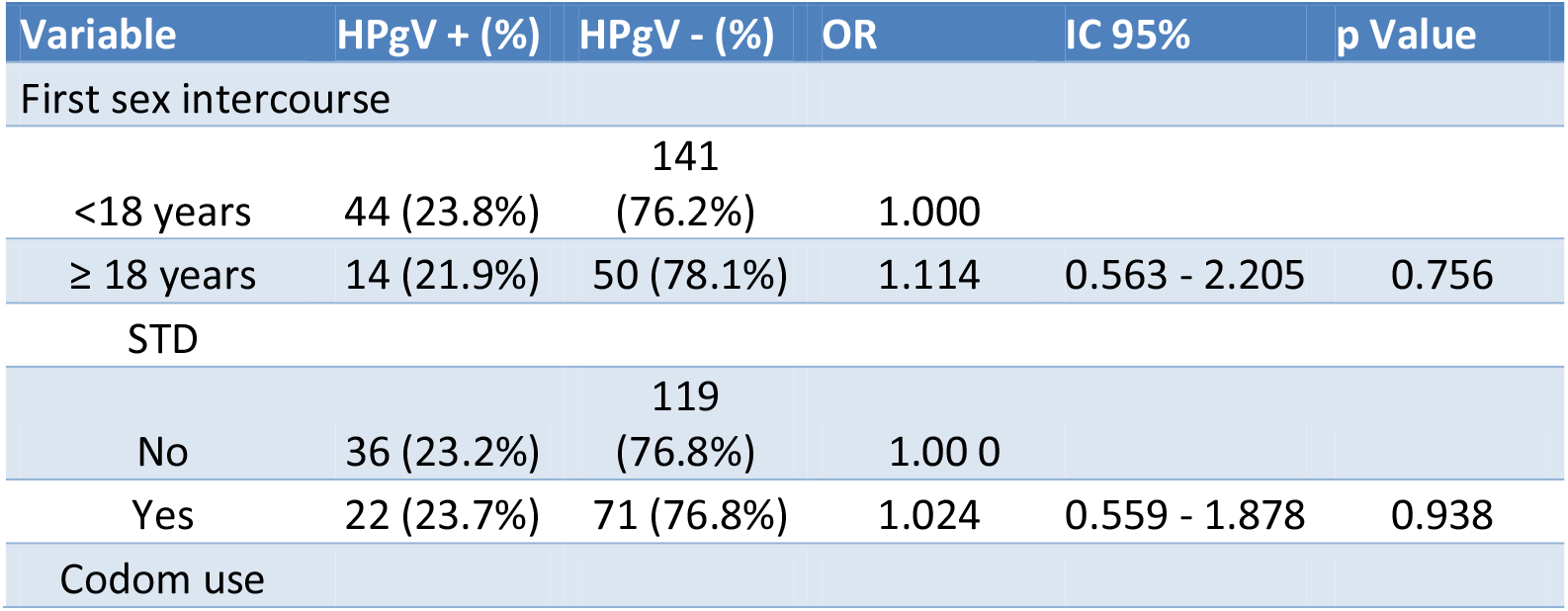
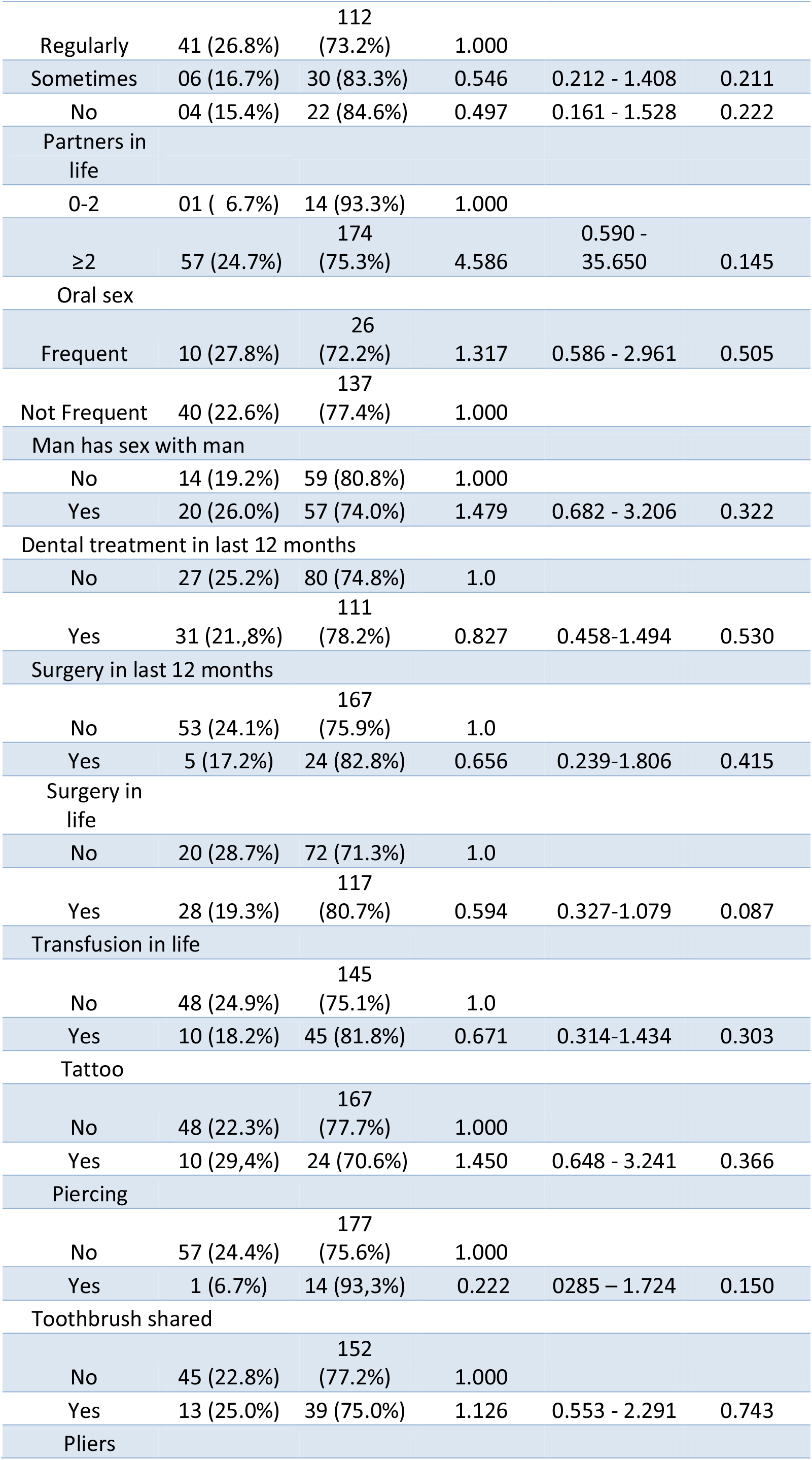
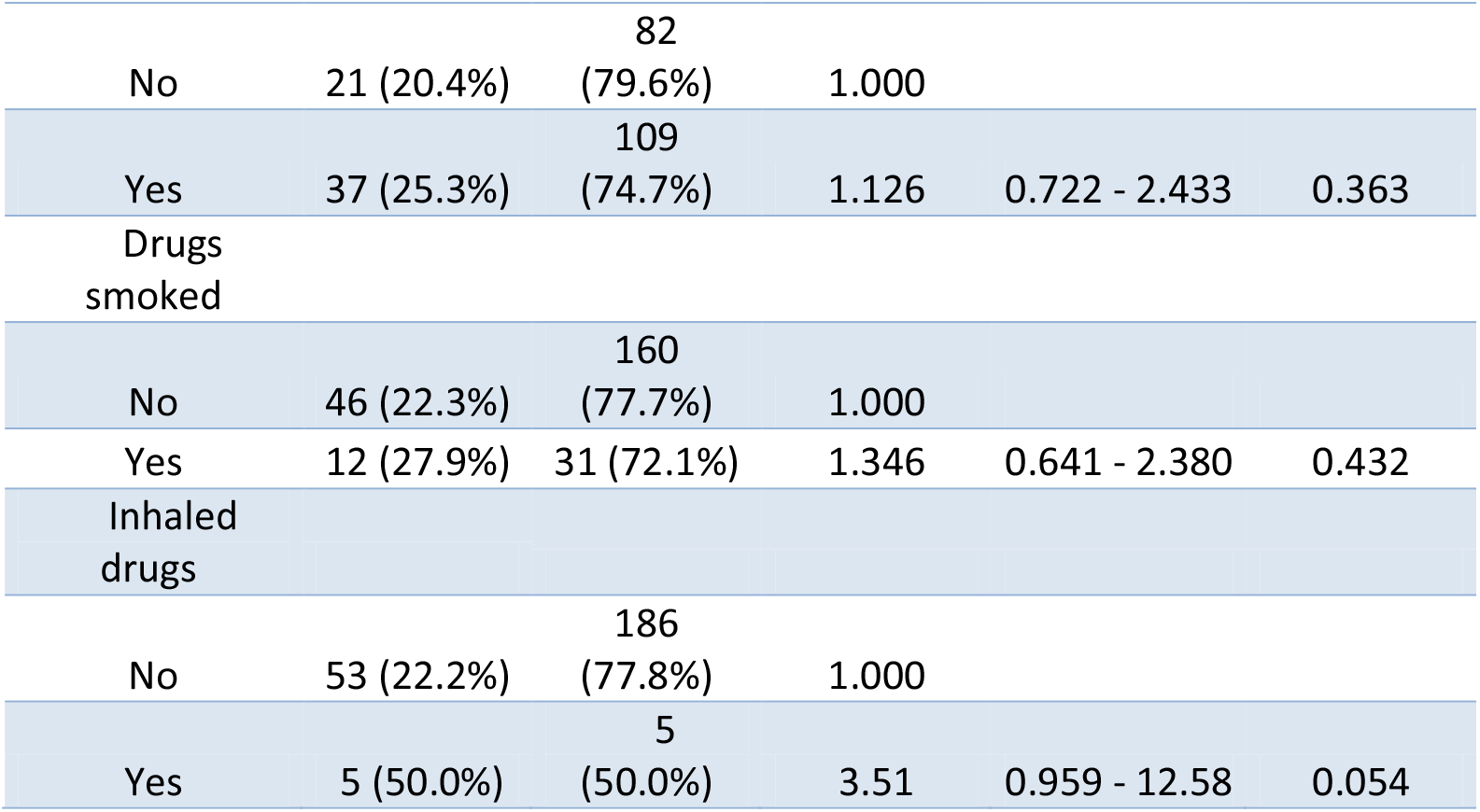
Univariate Analysis of the Association of the Variables Related to Sexually behavior and Parenteral Transmission and HPgV Infection in people living with HIV.

In the logistic regression model, we began by including the variable with the lowest p-value from the bivariate analysis, which was age with p = 0.039. Subsequently, we added the remaining variables to the model in ascending order of their p-values, while ensuring that the age variable remained statistically significant throughout the model-building process.

The forward stepwise logistic regression analysis identified nine variables as predictors of HPGV presence: age, piercing, inhaled drug use, income, and history of surgery. The model demonstrated acceptable fit (χ^2^ = 24.7, df = 9, p = 0.003) and explained 14.3% of the variance in HPGV presence (Nagelkerke R^2^ = 0.143). Omnibus likelihood ratio tests revealed significant associations between HPGV presence and age (χ^2^ = 7.51, df = 3, p = 0.057), piercing (χ^2^ = 8.45, df = 1, p = 0.004) and inhaled drug use (χ^2^ = 6.56, df = 1, p = 0.010),. History of surgery was not significantly associated with HPGV presence in the multivariable model (χ^2^ = 2.41, df = 2, p = 0.300).

Table 3 presents the estimated coefficients, standard errors, odds ratios, and 95% confidence intervals for each variable in the final model. Notably, compared to the youngest age group, older age groups were significantly more likely to have HPGV present. Conversely, individuals with piercings had a significantly lower odds of HPGV presence compared to those without piercings.

**Table 3.**
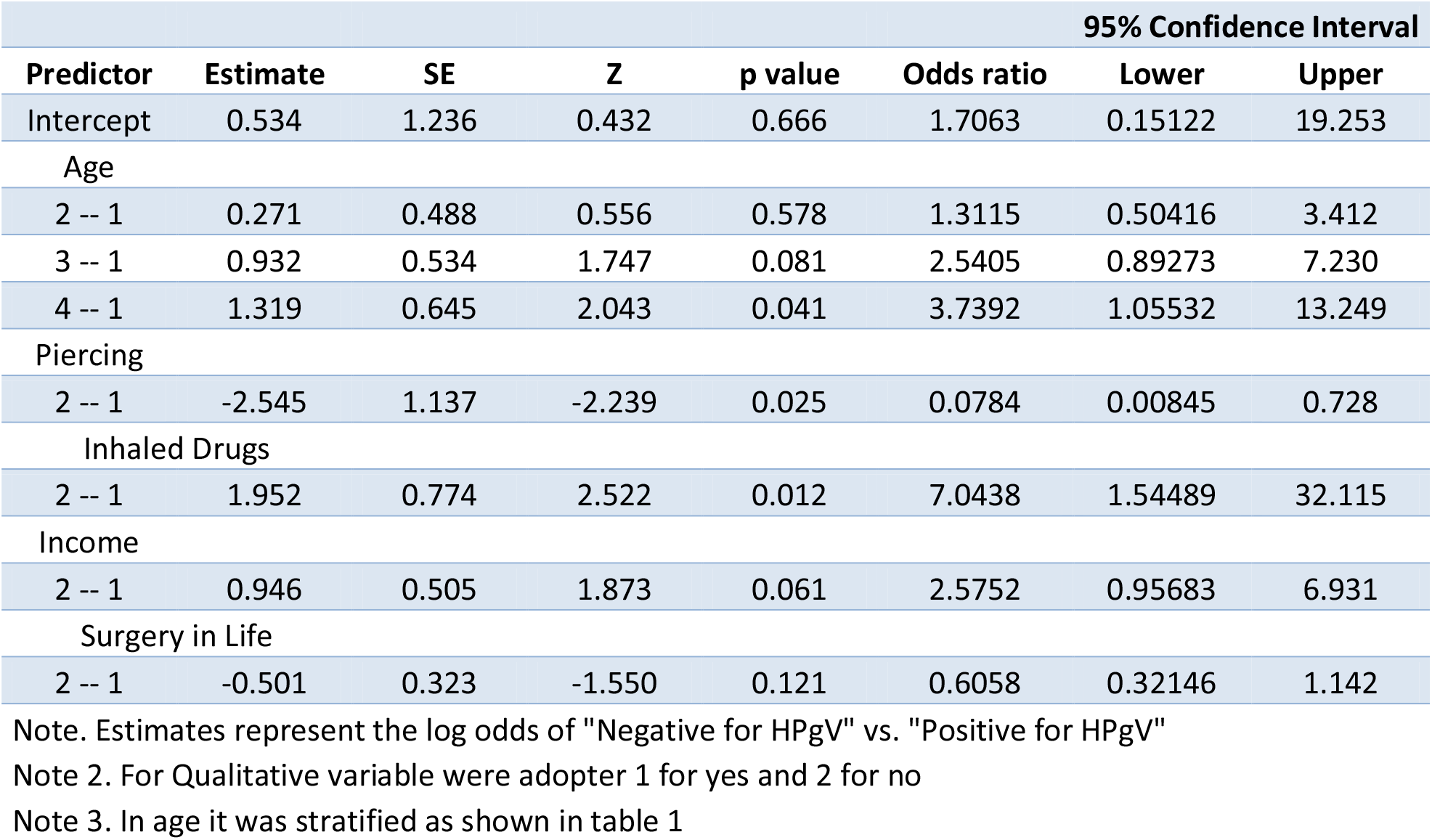
Logistic Regression Results for Predictors of HPGV Presence.

## DISCUSSION

Scholars have identified HPgV as a globally prevalent virus with a quiescent nature, persistently endeavoring to occupy suitable ecological niches. Nevertheless, investigations have consistently indicated that HPgV exhibits intricate interactions with HIV in co-infected individuals, implying a more multifaceted role for this vírus

Our results showed a high prevalence rate of 23.3% of HPgV RNA in PLWHIV, reminiscent of other studies that reported prevalence of 23.4% in Yunnan province in China (9), 33 % in Mexico (6), 19.6% in Cape Verde, Africa (15) and 27.3% in Turkey (8). In Brazil, Mota et al. (2016) observed a prevalence rate of 32% in PLWHIV in the extreme South, exceeding the prevalence found in our study. In contrast, the study carried out by Miranda et al. (2017) in the North region showed a prevalence of 17%.These comparisons highlight the wide geographic distribution and varying prevalence rates of HIV/HPgV co-infection across different regions and countries. These findings suggest that factors such as sociodemographic characteristics, risk behaviors, and access to healthcare may be influencing the epidemiology of this co-infection in diverse geographic contexts.

Here we found that individuals over 50 years old were more likely to have HIV/HPgV coinfection, suggesting that exposure increases with age. This contrasts with other studies, which reported a higher coinfection rate among younger patients (9; 15), or found no association with age (8). These inconsistencies may stem from differences in study populations, sampling methods, analytical approaches, or even geographical and cultural factors that could influence transmission dynamics and disease progression across diverse settings.

Our analysis unexpectedly showed a lower likelihood of HPgV in individuals with piercings. However, this doesn’t necessarily mean piercings offer protection. This could be due to other factors like differing health behaviors or social networks that influence HPgV exposure (16; 17). It’s also possible that those with piercings are more likely to engage in risk-taking behaviors, potentially increasing their exposure to HPgV (18; 19). Importantly, research has shown a clear link between body piercings and the transmission of similar viruses like HBV and HCV (16; 17). Therefore, we need further research to understand this complex relationship and should be cautious about assuming any protective effect from piercings.

Our data revealed an association between HIV/HPgV co-infection and the use of inhaled drugs. Users often share these drugs, which can come into contact with blood or other bodily fluids in the nose and/or mouth, serving as a potential source of transmission (20; 21).

It is interesting to note that 23.8% (44/185) of PLWHA who began their sexual activity under the age of 18 tested positive for HPgV, compared to 21.9% (14/64) in those who began their sexual activity over the age of 18. However, despite this difference in positivity rates, we did not find a statistically significant association between the age of onset of sexual activity and the presence of HPgV. This suggests that, although there is a disparity in infection rates between the two groups, other factors may be influencing the presence of the virus.

The findings regarding variables related to condom use did not align with expectations and differed from other studies. Additionally, we lack information on condom use at the time of serological conversion. Similarly, we found no statistically significant association between infection with HPgV and the variables concerning men who have sex with men and having multiple partners. This contrasts with studies that identified an association (22; 23).

Our study has some limitations stemming from its cross-sectional design, which does not allow us to verify the temporal sequence and thus limits the interpretation of our results. Furthermore, HPgV RNA positivity indicates current infection, but does not provide information about virus clearance. Therefore, some individuals considered negative for HPgV may have had a past infection. We were also unable to assess the CD4+ lymphocyte count to verify the potential beneficial effect of HPgV on HIV infection because all patients were on ART. Additionally, we could not establish the phase of HIV infection, which influences the CD4+ count.

This pioneering investigation in Northeast Brazil reveals a concerning 23.3% prevalence of HPgV among people living with HIV, emphasizing the potential burden of this co-infection across the Americas. Our findings identify age over 50 and, notably, the use of inhaled drugs as significant risk factors, suggesting transmission routes beyond traditional modes. This underscores the need to reassess and expand current prevention and harm reduction strategies. Further research is crucial to unravel the complex interplay between HPgV and HIV, ultimately guiding evidence-based policies and programs to effectively mitigate the burden of these intertwined viral diseases across diverse populations.

This study was financed in part by the Coordenação de Aperfeiçoamento de Pessoal de Nível Superior -Brazil (CAPES)-. Finance Code 001.

## Data Availability

All data produced in the present study are available upon reasonable request to the authors

